# Investigating Differential Changes in Cardiovascular Mortality by Documented diabetes in the United States Before and During the COVID-19 Pandemic: A Comparative Time-Series Analysis

**DOI:** 10.64898/2026.11.12.26367977

**Authors:** Muhammad Usman Ghani, Haeyoon Chang, Shofikur Shuhag, Sharat Kamath, Shahzaib Chughtai, Mohammad Alfrad Nobel Bhuiyan, Carlos J. Rodriguez

**Affiliations:** Department of Medicine, Covenant HealthCare College of Medicine at Central Michigan University, Mount Pleasant, Michigan, USA; School of Health Sciences, Central Michigan University, Mount Pleasant, Michigan, USA; Department of Medicine, Albert Einstein College of Medicine, Bronx, New York, USA; Department of Epidemiology and Population Health, Albert Einstein College of Medicine, Bronx, New York, USA; Department of Medicine, Henry Ford St. John Hospital, Detroit, Michigan, USA; Department of Medicine, Corewell Health, Grand Rapids, Michigan, USA; Department of Internal Medicine, Valley Health System, Winchester Medical Center, Winchester, Virginia, USA; Department of Medicine, Louisiana State University Health Sciences Center, Shreveport, Louisiana, USA; Department of Pathology and Translational Pathobiology, Louisiana State University Health Sciences Center, Shreveport, Louisiana, USA

**Keywords:** Cardiovascular mortality, Diabetes, COVID-19, Difference-in-differences, Preventive Cardiology

## Abstract

**Background:** Whether diabetes modified pandemic-related changes in cardiovascular disease (CVD) mortality remains uncertain. We examined differential changes in US CVD mortality among deaths with versus without diabetes documented on the death certificate.

**Methods:** We analyzed CDC WONDER Multiple Cause of Death data from 2005 to 2024, identifying deaths with CVD as the underlying cause among adults aged ≥25 years. Cohorts were CVD deaths with diabetes documented as a contributing cause (CVD+DM) and CVD deaths without documented diabetes. Because pre-2020 trajectories differed, the primary comparison of age-adjusted rates used a comparative interrupted time-series model allowing cohort-specific pre-2020 trends. Difference-in-differences estimates (2015–2019 vs 2020–2024) were descriptive.

**Results:** CVD+DM mortality declined from 43.1 per 100,000 in 2005 to 34.3 in 2019 and was 38.4 in 2024; mortality without documented diabetes declined from 398.7 to 296.6 and was 289.8. In the primary analysis, CVD+DM mortality showed a 13.1% greater relative level increase at 2020 (95% CI, 10.4%–15.9%), with no significant differential change in slope thereafter. Descriptively, the ratio of rate ratios was 1.16, and the absolute difference-in-differences was 5.0 per 100,000.

**Conclusions:** After accounting for cohort-specific pre-2020 trends, CVD mortality with documented diabetes showed a greater relative increase at the 2020 transition. These findings describe differential mortality patterns rather than a causal effect of diabetes or the pandemic.

**WHAT IS NEW?:**

- Cardiovascular disease mortality with documented diabetes showed a 13% greater relative level increase at the 2020 transition than cardiovascular disease mortality without documented diabetes, after accounting for pre-2020 trends.
- The difference was concentrated in the level change around 2020, with no significant differential change in slope thereafter.
- Findings were directionally consistent across analyses using alternative baseline periods, breakpoints, and restriction to type 2 diabetes.

**WHAT ARE THE CLINICAL IMPLICATIONS?:**

- The findings suggest that people with coexisting cardiovascular disease and diabetes may warrant particular attention during periods of major health system disruption, including efforts to maintain continuity of cardiometabolic care.
- Because the analysis is based on death-certificate documentation, the findings should be interpreted as a population-level mortality signal rather than evidence that diabetes caused the observed mortality difference.

## INTRODUCTION

Multiple national analyses have reported excess cardiovascular mortality during the COVID-19 pandemic period, with disproportionate increases among racial and ethnic minority populations.^1–3^ The drivers of these trends are likely multifactorial, including direct effects of SARS-CoV-2 infection, delayed care, healthcare system strain, and worsening cardiometabolic risk profiles.^4,5^

Improvements in prevention and acute care of cardiovascular disease (CVD) in the US were associated with declining mortality over several years, with a decline of 8.9% between 2010 and 2019. Overall US CVD mortality rates returned to near 2010 levels with over 228,000 excess CVD deaths.^6^ For example, the largest increases were observed in hypertensive disease, heart failure, and arrhythmias.^7,8^ Increases were also observed in ischemic heart disease (11% relative increase) and hypertensive disease (17% relative increase) during the initial pandemic surge.^5^ These trends highlight that the pandemic disrupted previously improving cardiovascular mortality trends. Consistent with these trends, an analysis of 2019 to 2021 Illinois Vital Records showed increases in diabetes and cardiovascular-related mortality during the pandemic, including deaths unrelated to COVID-19 infection.^9^

In this context, it is critical to consider the role of cardiometabolic conditions, particularly diabetes. Diabetes is a major contributor to cardiovascular mortality risk and may be associated with greater increases in mortality during the pandemic-period.^10,11^ Focusing on diabetes-related cardiovascular mortality, Bashar et al. found that mortality declined by 29.2% between 1999 and 2019.^10^ However, mirroring overall CVD mortality, this trend was sharply reversed when diabetes-relative cardiovascular mortality rates increased by 15.5% in 2020 alone, corresponding to an estimated 16,009 excess deaths.^10^ In a contemporary evaluation of type 2 diabetes mellitus– related mortality trends, Ahmed et al. found concerning shifts in US diabetes-related mortality from 1999 to 2023 that suggest the improvements observed in earlier decades may be plateauing or reversing in the setting of worsening cardiometabolic risk factors and pandemic-related disruptions.^11^

Important gaps remain in understanding how diabetes is associated with cardiovascular mortality trends during the COVID-19 pandemic. Recent studies have documented concerning trends in cardiovascular mortality during the pandemic period, but critical methodological limitations prevent causal inference about whether cardiovascular mortality trends differed according to documented diabetes involvement. Jain et al. examined 1999 to 2019 CDC WONDER data on diabetes-related cardiovascular mortality, finding a similar reversal of declining trends from 2014 to 2019. Significant demographic and geographic disparities were elucidated. Males and rural counties experienced the sharpest increases.^12^

To address this gap, we assessed whether cardiovascular mortality trends differed between deaths with and without documented diabetes around the onset of the COVID-19 pandemic. Using a comparative cohort framework, we compared changes in mortality over time between these groups on both absolute and relative scales, and accounted for each cohort’s own pre-pandemic trend, to characterise differential mortality patterns rather than to infer a causal effect of diabetes.

## METHODS

### Data Source and Study Population

We obtained age-adjusted CVD mortality data from the Centers for Disease Control and Prevention Wide-ranging Online Data for Epidemiologic Research (CDC WONDER) database^13^.

Data were extracted from two complementary final files: the 1999–2020 Multiple Cause of Death file, and the 2018–2024 Multiple Cause of Death by Single Race file. We compared overlapping years (2018–2020) across the two files and confirmed concordant national counts for the study definitions. The final analytic series used data from the 1999-2020 file for 2005-2017 and the 2018-2024 Single Race file for 2018-2024. For the overlapping years 2018–2020, we compared age-specific death counts and population denominators between the 1999–2020 and 2018–2024 files across all seven age groups used in the analysis. Death counts and population denominators were identical in all 21 year-age cells, yielding identical age-specific and age-adjusted rates. We therefore used the 1999–2020 file for 2005–2017 and the 2018–2024 file for 2018–2024. Detailed CDC WONDER query specifications and cohort construction are provided in the **Supplemental Methods**. All data was accessed on September 3, 2026.

We defined two study cohorts using International Classification of Diseases, Tenth Revision (ICD-10) underlying cause-of-death codes. The CVD deaths with documented diabetes (CVD+DM) cohort included decedents with CVD as the underlying cause (ICD-10 codes I00– I99) and diabetes mellitus listed anywhere as a contributing cause on the death certificate (ICD-10 codes E10–E14). The CVD deaths without documented diabetes (CVD without documented diabetes) cohort included decedents with CVD as the underlying cause without diabetes listed as a contributing condition. This approach allowed us to compare pandemic-era mortality patterns between CVD deaths with versus without documented diabetes involvement, recognizing that these death certificate-based cohorts reflect mortality patterns rather than population-level disease prevalence or risk.

The study period spanned 2005–2024, enabling examination of long-term pre-pandemic trends (2005–2019) and pandemic-era changes (2020-2024). The descriptive difference-in-differences (DID) comparison used the pre-pandemic period 2015–2019 versus the pandemic period 2020– 2024. Throughout this paper, “pre-pandemic” refers to 2015–2019 and “pandemic” to 2020– 2024, with the year 2020 included in the pandemic period.

### Age-Adjusted Mortality Rates

For the selected age range, standard-population weights were calculated as the 2000 U.S. standard population for each included age group divided by the total standard population across all included age groups, such that the weights summed to 1. We calculated age-adjusted mortality rates (AAMR) for adults aged ≥25 years using the direct method with the 2000 US standard population as the reference. Rates were computed overall for each cohort. Confidence intervals for age-adjusted rates were calculated using the Fay-Feuer method^14^, a gamma distribution approximation recommended by CDC for age-adjusted rates. We suppressed rates based on fewer than 10 deaths per CDC data-use restrictions and marked unreliable estimates when based on fewer than 20 deaths.

### Statistical Analysis

Annual age-adjusted mortality rates were first examined descriptively for each cohort. We calculated absolute and relative difference-in-differences (DID) estimates as descriptive summaries of changes in CVD mortality between the pre-pandemic (2015–2019) and pandemic (2020–2024) periods.^15,16^ For each cohort, period-specific rates were calculated as the arithmetic mean of the annual AAMRs for 2015–2019 and 2020–2024, respectively. The DID estimator was calculated as:

#### DID Estimate

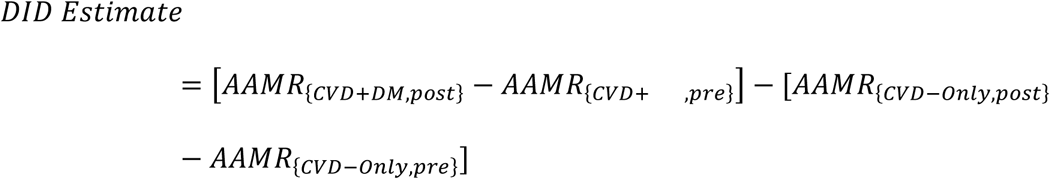

The relative DID was calculated as the ratio of the post-to-pre rate ratio in the CVD+DM cohort to the corresponding post-to-pre rate ratio in the cohort without documented diabetes, where “pre” refers to 2015–2019 and “post” refers to 2020–2024. A positive DID estimate indicates that CVD mortality with documented diabetes increased more, or declined less, than CVD mortality without documented diabetes during the pandemic period. The absolute and relative DID estimates are reported as descriptive measures without a formal significance test. Because the two cohorts differed substantially in baseline mortality rates, the relative scale was emphasized for comparative interpretation, while absolute estimates were retained as complementary measures.

Because the cohorts followed different pre-2020 trajectories, the primary trend-adjusted comparative analysis used a comparative interrupted time-series model of the annual ratio of age-adjusted mortality rates between the two cohorts on the log scale. The model included terms for the pre-2020 trend, the level change at 2020, and the change in slope after 2020. The level term estimated the relative change at 2020 after accounting for the preceding trend, while the post-2020 term estimated the subsequent change in slope. Standard errors accounted for potential correlation between adjacent annual observations. DID estimates were interpreted as descriptive pre/post summaries, whereas CITS estimates were used to characterize trend-adjusted temporal differences without implying causal effects.

### Parallel Trends Assessment

The validity of DID estimates relies on similarity of pre-pandemic trends between the comparison cohorts.^15^ We tested this assumption by fitting separate linear regression models to pre-pandemic AAMR (2005–2019) for each cohort and comparing annual slopes using interaction terms. We also examined whether trends converged during the immediate pre-pandemic period (2015–2019) that served as the DID baseline.

### Segmented Regression Analysis

As a supporting analysis, we used separate segmented regressions for each cohort to characterize within-cohort level and slope changes at the pre-specified 2020 breakpoint.^17^ For each cohort, we estimated:

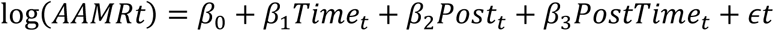

where *Time*_t_ = *Year* − 2005, *Post*_t_ = 1 for years 2020–2024 and 0 otherwise, and *PostTime*_t_ = max(0, *Year* − 2020). In this parameterization, *β*_1_represents the pre-2020 annual trend, *β*_2_ represents the immediate level change at 2020 relative to the projected pre-2020 trajectory, and *β*_3_ represents the change in annual slope after 2020. Annual percent changes were calculated as 100 × (*e*^β^ − 1).

### Robustness Checks

We conducted several sensitivity analyses, each designed to evaluate a specific threat to the validity of our primary estimate. First, because interrupted time-series estimates can be sensitive to the placement of the breakpoint, we re-estimated the CITS model using alternative breakpoints in 2019 and 2021 to assess whether the observed pattern relied on the 2020 inflection point. The same model specification was used, with standard errors adjusted for autocorrelation between adjacent annual observations. Second, because pre-pandemic trends were not fully parallel across the full baseline, we re-estimated the descriptive difference-in-differences using a shorter, more recent baseline (2017-2019) to determine whether the estimate was sensitive to the choice of baseline window. Third, because diabetes subtype may be misclassified on death certificates, we restricted the contributing-cause definition to type 2 diabetes (E11) to evaluate whether the findings depended on the broader E10–E14 definition. Finally, we examined pre-pandemic trend differences over the full pre-pandemic period (2005–2019).

All analyses were conducted using Python 3.12.10. Analysis code is available upon request to ensure reproducibility. We reported our findings in compliance with the Strengthening the Reporting of Observational Studies in Epidemiology (STROBE) guidelines.^18^

## RESULTS

### Population Characteristics

We observed 1,785,029 cardiovascular disease (CVD) deaths involving documented diabetes in the United States between 2005 and 2024 (**Table 1**). In comparison, 15,145,798 CVD deaths occurred without documented diabetes. Among CVD deaths involving documented diabetes, males accounted for 55.9% of deaths and females for 44.1%. Most deaths occurred among adults aged 65 years and older, particularly those aged 75–84 years (28.8%), followed by those aged 85 years and older (24.5%) and 65–74 years (23.7%).

**Table 1.** Characteristics of CVD Deaths With and Without Documented Diabetes, United States, 2005–2024.

| **Characteristic** | **CVD + documented diabetes, n (%)** | **CVD without documented diabetes, n (%)** |
|---|---|---|
| **Total deaths** | 1,785,029 | 15,145,798 |
| **Sex** |  |  |
| Male | 997,874 (55.9) | 7,555,903 (49.9) |
| Female | 787,155 (44.1) | 7,589,895 (50.1) |
| **Age group, y** |  |  |
| 25–34 | 4,882 (0.3) | 84,619 (0.6) |
| 35–44 | 26,475 (1.5) | 263,420 (1.7) |
| 45–54 | 106,427 (6.0) | 764,835 (5.0) |
| 55–64 | 272,842 (15.3) | 1,611,607 (10.6) |
| 65–74 | 422,594 (23.7) | 2,428,024 (16.0) |
| 75–84 | 514,257 (28.8) | 3,877,078 (25.6) |
| ≥85 | 437,552 (24.5) | 6,116,215 (40.4) |

Footnote: Values are n or n (%). CVD+DM denotes CVD deaths with diabetes documented as a contributing cause. CVD = cardiovascular disease; DM = diabetes mellitus.

### Overall Mortality Trends (2005–2024)

During 2005–2024, age-adjusted CVD mortality rates declined in both cohorts (***Figure 1, Supplemental Table S1***). The CVD deaths with documented diabetes cohort exhibited a pre-pandemic decline from 43.1 to 34.3 per 100,000 in 2005 and 2019, respectively (APC = −1.5% per year), followed by a higher pandemic-period level, averaging 39.9 per 100,000 during 2020– 2024, with a gradual decline to 38.4 per 100,000 in 2024. This pattern was consistent with an upward level shift at 2020 followed by continued gradual decline rather than a sustained upward trend. In contrast, the CVD deaths without documented diabetes cohort demonstrated a more pronounced pre-pandemic decline from 398.7 to 296.6 per 100,000 in 2005 and 2019, respectively, with continued decline to 289.8 per 100,000 in 2024.

**Figure 1.**
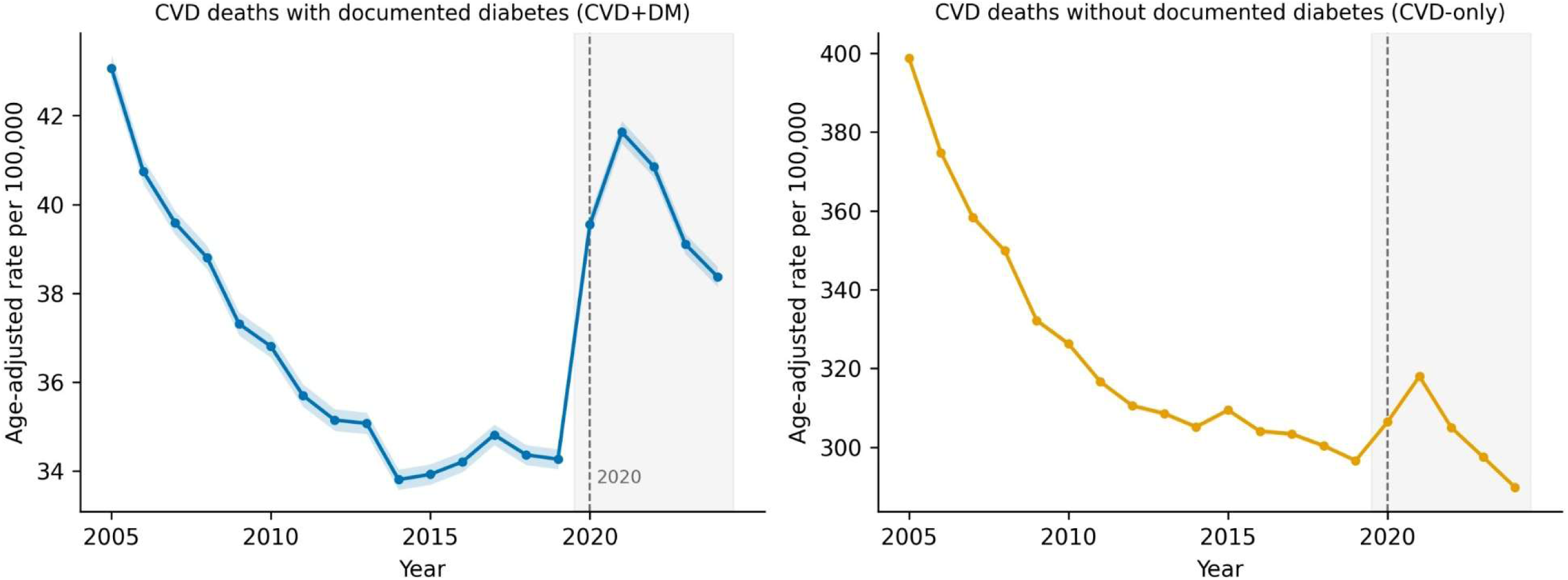
Age-Adjusted Cardiovascular Disease Mortality With and Without Documented Diabetes, United States, 2005–2024. The dashed vertical line indicates the 2020 transition; the shaded region denotes 2020–2024. Rates are age adjusted to the 2000 U.S. standard population and expressed per 100,000 adults aged ≥25 years. CVD = cardiovascular disease; DM = diabetes mellitus.

### Segmented Regression Trend Changes

As a supporting analysis, we used segmented regression analysis to examine whether mortality trends changed around the 2020 transition ***(Table 2)***. Neither cohort demonstrated a statistically significant change in slope after 2020. For the CVD+DM cohort, the change in post-2020 slope was not statistically significant (P=.69). Similarly, the CVD without documented diabetes cohort showed no significant change in post-2020 slope (P=.92). Thus, although mortality levels increased around 2020, there was no evidence that the subsequent long-term rate of decline changed significantly in either cohort.

**Table 2.**
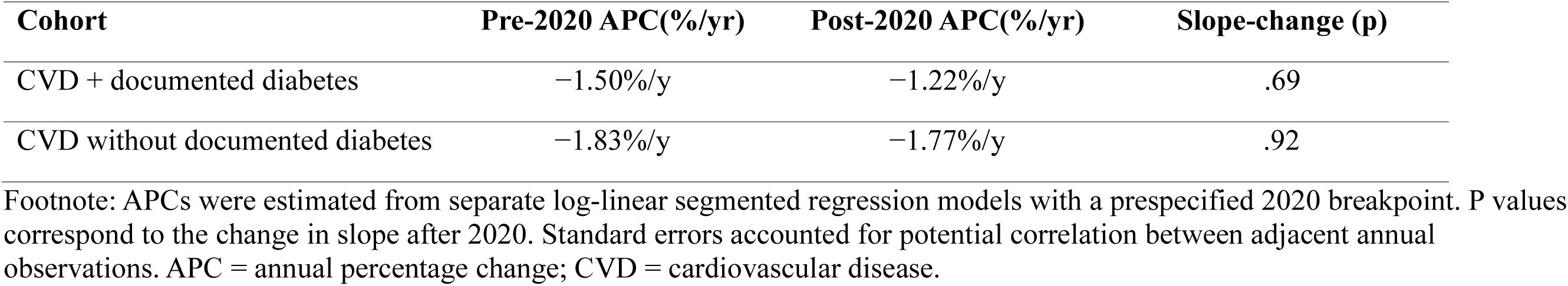
Segmented Regression of CVD Mortality Trends at the 2020 Breakpoint, United States, 2005–2024.

### Descriptive DID and Trend-Adjusted Comparative Interrupted Time-Series Analysis

As a descriptive comparison of changes between 2015–2019 and 2020–2024, CVD mortality with documented diabetes changed 16.1% more on the relative scale than CVD mortality without documented diabetes, corresponding to a ratio of rate ratios of 1.16 (***Table 3, Figure 2***). In the trend-adjusted comparative interrupted time-series analysis, the CVD+DM cohort demonstrated a 13.1% greater relative level increase in 2020 compared with the cohort without documented diabetes (95% CI, 10.4%–15.9%; P< .001). There was no significant differential change in slope thereafter (0.23% per year; 95% CI, −0.23 to 0.69; P=.33). The absolute difference-in-differences estimate was 5.0 deaths per 100,000. The difference between the absolute and relative estimates reflects the substantially different baseline mortality rates of the two cohorts. Findings were directionally similar in sensitivity analyses using a shorter 2017–2019 baseline period, a 2021 breakpoint, and restriction to type 2 diabetes (E11). In the CITS sensitivity analyses, the immediate relative level change remained significant with the 2021 breakpoint and E11 restriction, whereas the estimate was attenuated with a 2019 breakpoint and its confidence interval included the null (***Table 3***).

**Figure 2.**
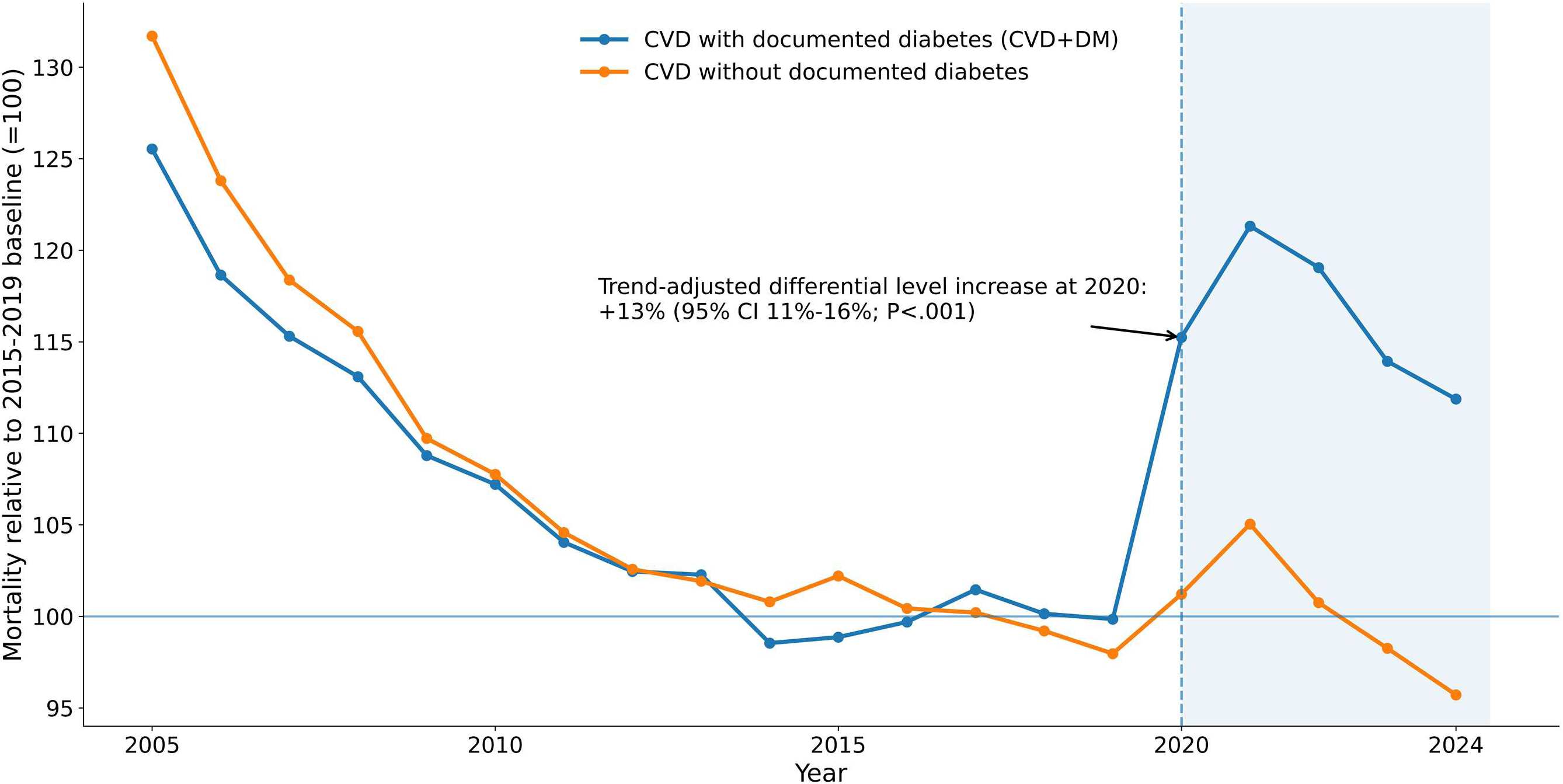
Relative Cardiovascular Disease Mortality With and Without Documented Diabetes, Indexed to the 2015–2019 Baseline, United States, 2005–2024. Values are indexed to the mean age-adjusted mortality rate during 2015–2019 for each cohort (=100). The dashed vertical line indicates the 2020 transition; the shaded region denotes 2020–2024. The annotated estimate represents the trend-adjusted differential level change from the comparative interrupted time-series analysis. CITS = comparative interrupted time-series; CVD = cardiovascular disease; DM = diabetes mellitus.

**Table 3.** Descriptive Difference-in-Differences and Comparative Interrupted Time-Series Analysis of CVD Mortality With and Without Documented Diabetes.

| **Analysis** | **Absolute DID, per 100,000** | **Relative DID, ratio of rate ratios** | **CITS immediate differential level change, % (95% CI)** | **P** |
|---|---|---|---|---|
| Primary: E10–E14, 2020 breakpoint | +5.0 | 1.16 | 13.1 (10.4 to 15.9) | <.001 |
| Sensitivity: 2019 breakpoint | +6.7 | 1.14 | 6.6 (−0.7 to 14.4) | .074 |
| Sensitivity: 2021 breakpoint | +5.6 | 1.14 | 11.0 (4.2 to 18.2) | .003 |
| Sensitivity: type 2 diabetes (E11) | +2.3 | 1.36 | 19.4 (5.7 to 34.9) | .007 |
| Sensitivity: shorter baseline (2017–2019) | +2.2 | 1.14 | — | — |

Footnote: DID estimates are presented as descriptive summaries without formal significance testing. Comparative interrupted time-series models used the annual log rate ratio of CVD mortality with versus without documented diabetes and HAC/Newey-West standard errors with a lag of 1 year. In the primary analysis, there was no significant differential change in slope after 2020 (0.23% per year; 95% CI, −0.27% to 0.73%; P=.34). DID= difference-in-differences; CITS=comparative interrupted time-series.

### Robustness and Sensitivity Analyses

Pre-2020 trends differed between cohorts over the full 2005–2019 period, indicating that the conventional parallel-trends assumption was not fully satisfied. Differences were smaller during the more recent 2015–2019 period that served as the DID baseline. Accordingly, the DID estimates were interpreted as descriptive summaries, while the comparative interrupted time-series analysis accounted for cohort-specific pre-2020 trajectories.

Sensitivity analysis using a shortened pre-pandemic baseline (2017–2019) demonstrated a DID estimate with similar direction and magnitude, supporting the consistency of the descriptive findings despite baseline differences. Sensitivity analysis using alternative breakpoint specifications produced generally similar patterns. When testing alternative breakpoints at 2019 and 2021, slope changes remained non-significant and magnitude estimates were consistent, indicating that the estimates were similar under the alternative breakpoint specifications.

Further sensitivity analysis restricting diabetes to type 2 diabetes (E11) yielded a directionally consistent absolute DID estimate of 2.3 per 100,000.

## DISCUSSION

In this national analysis of cardiovascular mortality trends using CDC WONDER data, age-adjusted cardiovascular mortality among the CVD+DM cohort increased more during the pandemic era than rates among the cohort without documented diabetes. After accounting for each cohort’s own pre-2020 trend, the between-cohort difference was statistically significant, with a differential relative level increase of 13.1% at 2020 (95% CI, 10.4%–15.9%). On the absolute scale, the descriptive difference-in-differences estimate was 5.0 deaths per 100,000. The difference between the absolute and relative estimates reflects the substantial difference in baseline mortality rates between the two groups. We interpret these findings as descriptive of a differential pandemic-era pattern rather than a causal effect of diabetes, and our analysis reflects death-certificate documentation rather than population-level risk. Prior to 2020, age-adjusted cardiovascular mortality declined in both cohorts, followed by an upward shift around 2020 and subsequent decline. Ours findings were generally supported by sensitivity analyses using a 2021 breakpoint and restriction to type 2 diabetes, although the immediate level-change estimate was attenuated when the breakpoint was shifted to 2019, and its confidence interval included the null.

Our study found the divergence in long-term mortality trends between CVD deaths with and without diabetes involvement. Cardiovascular mortality among CVD deaths without documented diabetes declined substantially over the study period, consistent with the broader national decline in CVD mortality. In contrast, mortality among CVD deaths with documented diabetes declined less. This divergence suggests a continuing and relatively greater burden of cardiometabolic disease among decedents with documented diabetes, although our data cannot identify the factors underlying it.

The observed mortality patterns may reflect several overlapping converging mechanisms. Diabetes is associated with endothelial dysfunction, oxidative stress, systemic inflammation, dyslipidemia, and a prothrombotic state, accelerating atherosclerosis, impair vascular repair, and increase the risk of myocardial infarction, stroke, and heart failure. ^19–22^ Diabetes may also cause structural and metabolic changes in the myocardium (diabetic cardiomyopathy) that worsen outcomes following ischemic injury.^23,24^ In addition, impaired immune responses may contribute to more severe COVID-19 illness, while disruptions in glucose monitoring, medication management, and preventive care may have worsened metabolic control. These biological and healthcare-delivery factors provide plausible explanations for the observed mortality patterns. Population-based studies have consistently shown that individuals with diabetes experience substantially higher cardiovascular mortality compared with those without diabetes,^25–28^ and our findings extend this literature to the pandemic period.

The COVID-19 pandemic introduced major disruptions to healthcare delivery that likely influenced cardiovascular outcomes. Several national analyses have reported increases in cardiovascular mortality during the pandemic period, including deaths not directly attributed to SARS-CoV-2 infection.^9^ At the same time, declines in hospitalizations for acute myocardial infarction and other cardiovascular emergencies were observed during the early months of the pandemic, suggesting that many patients delayed or avoided seeking care.^29,30^ Reductions in outpatient visits, laboratory monitoring, and preventive services were also widely documented during this period.^31^ These disruptions may have had a greater impact on individuals with diabetes, whose disease management often depends on regular monitoring and medication adjustments. Interruptions in routine care could therefore contribute to worsening glycemic control and increased cardiovascular risk. In addition, individuals with diabetes were more likely to experience severe COVID-19 infection, which has been associated with myocardial injury, systemic inflammation, and thrombotic complications.^32^

### Public Health Implications

Our findings suggest several important implications for public health preparedness and clinical practice. Although our analysis did not evaluate specific interventions, the observed mortality patterns are consistent with the view that individuals with concurrent CVD and diabetes may represent a high-priority population during future public health emergencies. Maintaining continuity of chronic disease management during crises, and anticipating the compounded risks of multimorbidity, may be reasonable considerations for preparedness planning. Healthcare systems may benefit from contingency plans to maintain continuity of chronic disease management during crises, including telehealth infrastructure, home monitoring capabilities, and medication delivery systems. Public health messaging could explicitly address potential compounded risks of multiple chronic conditions, encouraging heightened precautions among populations with multimorbidity. Clinically, our results reinforce the importance of integrated cardiovascular-metabolic care. The interconnected nature of CVD and diabetes suggests that optimizing management of one condition may buffer risk from the other. Pandemic-era care disruptions may have disproportionately affected individuals with complex multimorbidity, highlighting the need for care models that provide coordinated rather than fragmented chronic disease management.

### Limitations

Several limitations should be considered. First, pre-2020 mortality trends differed between cohorts, so the parallel-trends assumption required for conventional DID interpretation was not fully satisfied. We therefore treated the absolute and relative DID estimates as descriptive summaries and used a comparative interrupted time-series model that allowed cohort-specific pre-2020 trends as the primary analysis. Results were directionally consistent using a shorter baseline and alternative breakpoint specifications, although effect estimates varied across analyses. Second, our study relies on death-certificate data and compares CVD deaths with versus without documented diabetes involvement rather than population-level mortality risk among individuals with and without diabetes. Diabetes may be underreported, differentially recorded, or misclassified by subtype; restricting the analysis to type 2 diabetes produced a similar direction of effect, though the magnitude differed. Consistent underreporting across the study period would be less likely to explain a differential change at 2020; however, increased diabetes documentation during the pandemic could have overestimated the relative increase in the CVD+DM cohort. Death certificates also do not provide information on glycemic control, disease duration, medication use, health care access, or other individual-level factors that may have influenced mortality. Third, we could not distinguish changes directly related to COVID-19 infection from indirect pandemic-era effects such as disruptions in care or changes in health behavior. The findings should therefore be interpreted as temporal associations around the 2020 transition rather than causal effects of the pandemic. Finally, changes in population-estimation methods beginning in 2021 may have affected absolute age-adjusted rate comparisons across the study period.

### Future Directions

Several research directions emerge from this analysis. First, patient-level studies examining specific mechanisms (acute COVID-19 severity versus chronic care disruptions) would clarify intervention targets. Second, analysis of medication utilization and healthcare engagement patterns could reveal modifiable contributors to excess mortality. Third, examination of post-pandemic recovery trajectories (2025 onward) will help determine whether observed excess mortality persists or resolves. Fourth, international comparisons could reveal whether healthcare system characteristics (universal vs. fragmented coverage, primary care accessibility) modified the diabetes-related pandemic impact.

## CONCLUSION

In this 20-year analysis of national mortality data, CVD mortality with documented diabetes demonstrated a greater relative increase at the 2020 transition than CVD mortality without documented diabetes after accounting for pre-2020 mortality trends. The trend-adjusted relative difference was approximately 13%, whereas the corresponding descriptive absolute difference-in-differences was 5.0 deaths per 100,000, reflecting the differential baseline mortality rates of the two groups. These findings are best interpreted as a descriptive temporal differential rather than a causal effect and suggest that the mortality increase around 2020 was proportionally greater among CVD deaths with documented diabetes. The results also highlight the importance of maintaining chronic disease management and continuity of care during major public health emergencies.

## Nonstandard Abbreviations and Acronyms

AAMR: age-adjusted mortality rate
APC: annual percentage change
CITS: comparative interrupted time-series
CVD+DM: cardiovascular disease deaths with diabetes documented as contributing cause
DID: differences-in-differences
DM: diabetes mellitus

## Data Availability Statement

The data underlying this study are publicly available from the CDC WONDER online database (https://wonder.cdc.gov). The specific query parameters used to extract the mortality data are provided in the Supplemental Material.

## Supplemental Material

Supplemental Methods

Table S1

